# Forces Applied on the Glottis During Endotracheal Intubation: Effect of Technique, Stylet, and Experience – A Manikin-based study

**DOI:** 10.64898/2026.03.05.26347753

**Authors:** Louis Morisson, Arnaud Latreille, Mathias Pietrancosta, Katia Djerroud, Issam Tanoubi, Thomas Hemmerling, Pascal Laferrière-Langlois

**Author notes:** **Corresponding author:** Pascal Laferrière-Langlois, M.D., M.Sc. **Address:** 5415 Boulevard de l’Assomption, Montréal, Québec, Canada, H1T 2M4, **Email:**.

## Abstract

**Purpose:** To quantify and compare the peak force applied on the glottis during endotracheal intubation across five laryngoscopy techniques, two intubation conditions (standard and simulated laryngospasm), and two operator experience levels, and to assess the effects of stylet use and operator anthropometric characteristics on applied force.

**Methods:** This prospective, manikin-based experimental study enrolled 50 operators (30 experienced, 20 less experienced). Each performed endotracheal intubation using five techniques: direct laryngoscopy and videolaryngoscopy with a Macintosh blade, each with and without stylet, and videolaryngoscopy with a hyperangulated blade with stylet. A calibrated force sensor positioned at the glottis measured peak forces during standard and simulated laryngospasm conditions. Non-parametric statistical methods were used (Mann-Whitney U, Wilcoxon signed-rank, Friedman tests); effect sizes are reported as rank-biserial correlations.

**Results:** Across all techniques, median glottic forces ranged from 4.8 N (IQR: 3.3–6.5) for videolaryngoscopy without stylet to 11.1 N (IQR: 7.5–14.5) for direct laryngoscopy with stylet under standard conditions. No significant differences in applied force were observed between experienced and less experienced operators for any technique–condition combination (all adjusted p = 1.0; |r| ≤ 0.27). Stylet use significantly increased glottic force across all conditions and groups (median increases 3.4–7.3 N; all p < 0.001; rank-biserial r ≥ 0.75). Videolaryngoscopy with a Macintosh blade produced significantly lower forces than hyperangulated videolaryngoscopy under standard conditions (adjusted p = 0.049). Neither grip strength nor hand size correlated with applied force.

**Conclusion:** Glottic force during endotracheal intubation is determined primarily by technique and stylet use, not operator experience or anthropometrics. Stylet use is the single largest modifiable contributor to glottic force. These findings have implications for device selection, clinical training, and strategies to minimize airway trauma during intubation.

**IMPLICATION STATEMENT:** This manikin-based study quantifies glottic forces during endotracheal intubation across laryngoscopy techniques, stylet use, and operator experience levels, providing the first comprehensive characterization of right-hand forces transmitted through the tube. Stylet use consistently and substantially increases glottic force regardless of technique or operator experience, informing device selection and training strategies to minimize airway trauma.

## BACKGROUND

Endotracheal intubation is a fundamental procedure in anesthesia, emergency medicine, and critical care, performed millions of times annually worldwide. Despite its routine nature, intubation-related injuries remain a recognized complication with a broad clinical spectrum. Iatrogenic tracheobronchial injuries occur in approximately 0.005% of all endotracheal intubations, with rates reaching 0.5% for double-lumen tubes^1^.Dental injuries occur in approximately 1 in 150 intubations, predominantly involving the maxillary central incisors^2,3^. Postoperative sore throat, one of the most common complaints, affects up to 60% of intubated patients, with mechanical trauma from forced laryngoscopy and stylet use being identified as key contributing factors.^4^ Arytenoid dislocation, though uncommon (incidence 0.01–0.1%), has been reported in up to 30% of patients referred for persistent hoarseness after laryngeal instrumentation.^5^

While forces exerted by the laryngoscope blade on airway tissues have been extensively studied, forces exerted by the endotracheal tube itself on the larynx have only recently been quantified in cadaveric models, demonstrating that larger tube diameters produce greater forces on the laryngotracheal complex.^6^ Research using cadaveric models has also demonstrated that direct forces up to 4.7 kg (46 Newtons [N]) could not dislocate the cricoarytenoid joint, suggesting that substantial mechanical force is required to cause such injuries.^7^ However, a critical knowledge gap exists: no comprehensive studies have quantified the forces routinely applied on the glottis during standard intubation procedures, nor have they established the maximum forces that operators are willing to exert during challenging clinical scenarios.

Previous studies using force sensors on laryngoscope blades have shown that videolaryngoscopy generally requires less lifting force to achieve glottic visualization than direct laryngoscopy. Hindman et al. reported Macintosh forces of 48.8 N compared to Airtraq forces of 10.4 N,^8^ and Cordovani et al. demonstrated lower peak forces with the GlideScope compared to the Macintosh laryngoscope.^9^ However, these measurements captured the force applied by the left hand on the laryngoscope blade against the tongue base and vallecula. The force applied by the right hand on the glottis itself, transmitted via the endotracheal tube and stylet during tube passage through the vocal cords, has not been specifically characterized. This distinction is clinically important as the laryngoscope exerts pressure in a different axis and on different structures than the right hand.

The use of an intubating stylet increases endotracheal tube rigidity, which may influence force transmission to laryngeal structures.^10^ While stylet use significantly improves first-attempt intubation success, as demonstrated in the STYLETO trial (78.2% vs. 71.5%, p = 0.01),^11^ the increased rigidity may reduce the tube’s ability to deflect upon contact with the glottis, potentially resulting in higher contact forces. Clinical expertise might also influence force application during intubation, as experienced operators develop refined technique efficiency through extensive practice.^12^ However, objective data quantifying force differences across experience levels are lacking.

This investigation was designed to provide the first comprehensive quantitative characterization of the force applied on the glottis during endotracheal intubation, specifically the force transmitted by the right hand via the tube and stylet. We sought to explore the effect of stylet use, operator experience level, and laryngoscopy technique on glottic force, using five technique combinations and two force conditions. This baseline data will inform current clinical practice, training protocols, and equipment development initiatives.

## HYPOTHESIS

We hypothesized that force application during intubation would vary significantly across the five intubation techniques. We also hypothesized that the use of a stylet would significantly increase the force applied on the glottis compared to intubation without a stylet, regardless of laryngoscopy technique or operator experience level. Finally, we hypothesized that experienced operators would apply lower forces during standard intubation compared to less experienced operators, with a significant interaction between experience level and technique type.

## METHODS

### Study Design

This prospective, manikin-based experimental study was conducted in the Department of Anesthesiology and Pain Medicine, Hôpital Maisonneuve-Rosemont (Montréal, Québec, Canada) to measure and compare the force applied on the glottis during endotracheal intubation using different laryngoscopy techniques. The study protocol was designed in accordance with the STROBE statement for reporting observational studies.^13^

### Ethics

This study was approved by our local ethics committee (Comité d’éthique de la recherche du CIUSSS de l’Est de l’Ile de Montréal, Montréal, Québec, Canada) on december 10^th^ 2025 (Project number: 2026-4206). All participants signed the informed consent form.

### Participants

A total of 50 operators were recruited into two groups: 30 experienced operators and 20 less experienced operators comprising anesthesia residents (post-graduate year [PGY] 1 and 2), medical students with at least one prior intubation attempt on a real patient, and respiratory therapists. Experienced operators were defined as attending physicians and residents at PGY 3 and above in anesthesiology, emergency medicine, or intensive care medicine.

### Intubation Techniques and Experimental Setup

Each participant performed endotracheal intubation using five distinct techniques:

1. Direct laryngoscopy without stylet
2. Direct laryngoscopy with stylet
3. Videolaryngoscopy (Macintosh blade) without stylet
4. Videolaryngoscopy (Macintosh blade) with stylet
5. Videolaryngoscopy (hyperangulated blade) with stylet

The sequence of intubation techniques was randomized per a pre-established randomization list. For each technique, participants performed the intubation under two experimental conditions: (1) a *standard force* condition, in which they were instructed to intubate as they would in routine clinical practice; and (2) a *simulated laryngospasm* condition, in which glottic entry was blocked via an external pressure mechanism that mimicked a laryngospasm, and operators were instructed to apply the maximum force they would consider acceptable to insert the endotracheal tube and restore oxygenation. Additionally, a subset of experienced operators (n = 10) performed a third *physical maximum force* condition, representing the absolute maximum force they could physically exert with the intubation equipment.

### Equipment

Direct laryngoscopy was performed using standard Macintosh laryngoscopes with size 3 blades. Videolaryngoscopy procedures utilized the McGrath MAC system (Aircraft Medical/Medtronic, UK) with two blade configurations: Macintosh-style blades (size 3) and hyperangulated X3 blades. Standard malleable intubating stylets were used for Macintosh-style blade intubation, shaped by the operator prior to intubation; a rigid stylet was used for hyperangulated blade intubation. Endotracheal tubes were size 7.5 mm internal diameter with high-volume, low-pressure cuffs.

### Force Measurement

The force applied on the glottis was measured using a calibrated force sensor (Digital Force Gauge M5-20; Mark-10 Corporation, NY, USA) positioned immediately below the vocal cords in a high-fidelity airway training manikin, capturing the force transmitted by the right hand via the endotracheal tube and stylet during tube passage (Figure 1). Force data were recorded continuously in Newtons (N) throughout each intubation attempt. The peak (maximum) force value recorded during each intubation was extracted as the primary outcome. Anthropometric measurements, including dominant hand grip strength (measured using a hand dynamometer in kilograms) and hand size (measured in centimetres), were collected for each participant.

**Figure 1.**
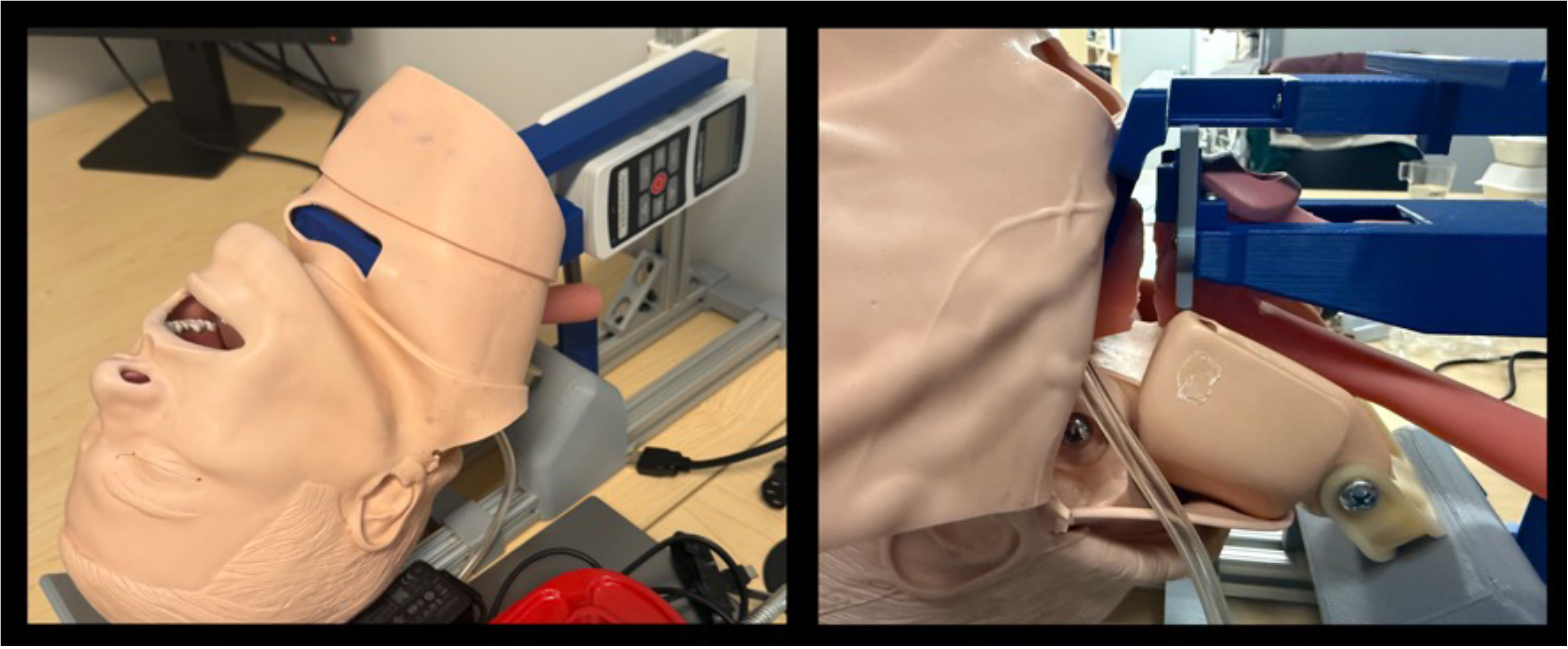
Experimental setup showing the airway training manikin with the force sensor positioned at the glottis. The Digital Force Gauge (Mark-10 M5-20) captures the force transmitted by the right hand via the endotracheal tube and stylet during tube passage through the vocal cords.

### Statistical Analysis

Normality of the force distributions was assessed using the Anderson-Darling test^14^ complemented by evaluation of skewness and kurtosis. The Anderson-Darling test rejected the null hypothesis of normality for 43% of the distributions tested at α = 0.05, and 52% of distributions exhibited high skewness (|skewness| > 1.0). Force distributions were consistently right-skewed, as expected for physiological force data bounded at zero. Consequently, non-parametric statistical methods were used throughout, and descriptive statistics are reported as median (IQR).

Between-group comparisons (experienced vs. less experienced) were performed using the Mann-Whitney U test^15^ for each technique–condition combination. The rank-biserial correlation (r) was calculated as the non-parametric effect size measure.^16^ Multiple comparison correction was applied using the Holm-Bonferroni step-down method across the 10 comparisons.^17^

The effect of the stylet on applied force was assessed using the Wilcoxon signed-rank test for paired samples,^18^ comparing intubation with stylet versus without stylet within each participant for both direct laryngoscopy and videolaryngoscopy with Macintosh blade. The matched-pairs rank-biserial correlation was used as the effect size measure. Analyses were conducted separately for experienced operators, less experienced operators, and the combined cohort.

Comparison across the three laryngoscopy techniques using a stylet was performed using the Friedman test for repeated measures.^19^ When the Friedman test was significant, post-hoc pairwise comparisons were conducted using the Conover test^20^ with Holm-Bonferroni correction. Correlations between anthropometric variables and applied force were assessed using Spearman rank correlation coefficients.^21^ All statistical tests were two-tailed with a significance level of α = 0.05. Effect sizes were interpreted according to conventional thresholds: |r| < 0.10 as negligible, 0.10–0.30 as small, 0.30–0.50 as medium, and > 0.50 as large.^22^

## RESULTS

### Description of Operators

The experienced group consisted of 24 anesthesiologists (80.0%) and 6 anesthesia residents (20.0%), with 23 participants (76.7%) performing more than 50 intubations per 3-month period and 7 (23.3%) performing 21 to 50. The less experienced group comprised 12 respiratory therapists (60.0%), 5 medical students (25.0%), 2 general surgeons (10.0%), and 1 anesthesia resident (5.0%), with 12 (60.0%) performing 0 to 5 intubations and 8 (40.0%) performing 5 to 20 intubations per 3-month period. Demographic characteristics are presented in Table 1.

**Table 1.** Participant characteristics.

| Characteristic | Experienced (n = 30) | Less Experienced (n = 20) | Total (N = 50) |
| --- | --- | --- | --- |
| ***Profession, n (%)*** |  |  |  |
| Anesthesiologists | 24 (80.0) | 0 | 24 (48.0) |
| Anesthesia residents | 6 (20.0) | 1 (5.0) | 7 (14.0) |
| Respiratory therapists | 0 | 12 (60.0) | 12 (24.0) |
| Medical students | 0 | 5 (25.0) | 5 (10.0) |
| General surgeons | 0 | 2 (10.0) | 2 (4.0) |
| ***Intubations per 3-month period, n (%)*** |  |  |  |
| >50 | 23 (76.7) | 0 | 23 (46.0) |
| 21–50 | 7 (23.3) | 0 | 7 (14.0) |
| 5–20 | 0 | 8 (40.0) | 8 (16.0) |
| 0–5 | 0 | 12 (60.0) | 12 (24.0) |
| ***Self-reported confidence (median, IQR)*** |  |  |  |
| Direct laryngoscopy | 9.0 (8.0–10.0) | 6.0 (5.0–7.0) | 8.0 (6.0–9.0) |
| Videolaryngoscopy | 10.0 (9.0–10.0) | 8.0 (7.5–9.0) | 9.0 (8.0–10.0) |

*IQR, interquartile range.*

Self-reported confidence for direct laryngoscopy was higher in experienced operators (median 9.0/10, IQR: 8.0–10.0) than less experienced operators (median 6.0/10, IQR: 5.0–7.0). Similarly, confidence for videolaryngoscopy was higher in the experienced group (median 10.0/10, IQR: 9.0–10.0) than the less experienced group (median 8.0/10, IQR: 7.5–9.0). The preferred intubation technique was videolaryngoscopy with a Macintosh blade for both groups.

### Descriptive Statistics

Peak glottic forces measured during intubation are summarized in Table 2. Across all techniques and both groups, median forces during standard intubation ranged from 4.8 N (IQR: 3.3–6.5) for videolaryngoscopy without stylet to 11.1 N (IQR: 7.5–14.5) for direct laryngoscopy with stylet (Figure 2 and 4 panels A-C). Under the simulated laryngospasm condition, median values ranged from 12.2 N (IQR: 9.6–16.3) for direct laryngoscopy without stylet to 20.2 N (IQR: 17.5–23.9) for hyperangulated videolaryngoscopy with stylet (Figure 3 and 4 panels D-F). In the experienced subgroup (n = 10) that performed the physical maximum force condition, the highest median force was observed with hyperangulated videolaryngoscopy with stylet at 31.1 N (IQR: 24.3–35.4).

**Table 2.** Peak glottic force (N) by technique, condition, and experience level. Values are expressed as median (IQR).

| Technique | All participants (N = 50) |  | Less experienced (N = 20) |  | Experienced (N = 30) |  |  |
| --- | --- | --- | --- | --- | --- | --- | --- |
|  | Standard | Clin. max | Standard | Clin. max | Standard | Clin. max | Phys. max (n = 10) |
| DL with stylet | 11.1 (7.5–14.5) | 20.0 (15.2–24.1) | 11.8 (7.4–15.1) | 20.4 (17.2–24.6) | 10.3 (7.6–14.5) | 19.3 (12.7–24.1) | 26.1 (22.8–32.6) |
| DL without stylet | 6.1 (3.9–9.8) | 12.2 (9.6–16.3) | 5.4 (3.6–7.4) | 14.4 (10.6–16.9) | 7.5 (4.3–11.7) | 11.3 (9.2–15.4) | 12.7 (8.3–17.6) |
| VL Macintosh with stylet | 8.7 (6.1–12.4) | 19.5 (15.4–24.0) | 8.5 (6.1–10.1) | 19.5 (17.2–23.0) | 8.7 (6.1–13.7) | 19.6 (14.7–24.5) | 24.7 (20.8–27.1) |
| VL Macintosh without stylet | 4.8 (3.3–6.5) | 12.3 (9.2–15.5) | 4.2 (3.2–5.0) | 11.3 (9.3–14.4) | 5.2 (3.6–7.3) | 12.7 (9.2–17.4) | 11.2 (7.5–16.4) |
| VL Hyperangulated with stylet | 10.3 (8.2–13.5) | 20.2 (17.5–23.9) | 10.0 (7.8–13.8) | 20.2 (17.7–23.2) | 10.3 (8.6–13.5) | 20.4 (17.5–23.9) | 31.1 (24.3–35.4) |

*DL, direct laryngoscopy; VL, videolaryngoscopy; Clin. max, clinical maximum force; Phys. max, physical maximum force (n = 10).*

**Figure 2.**
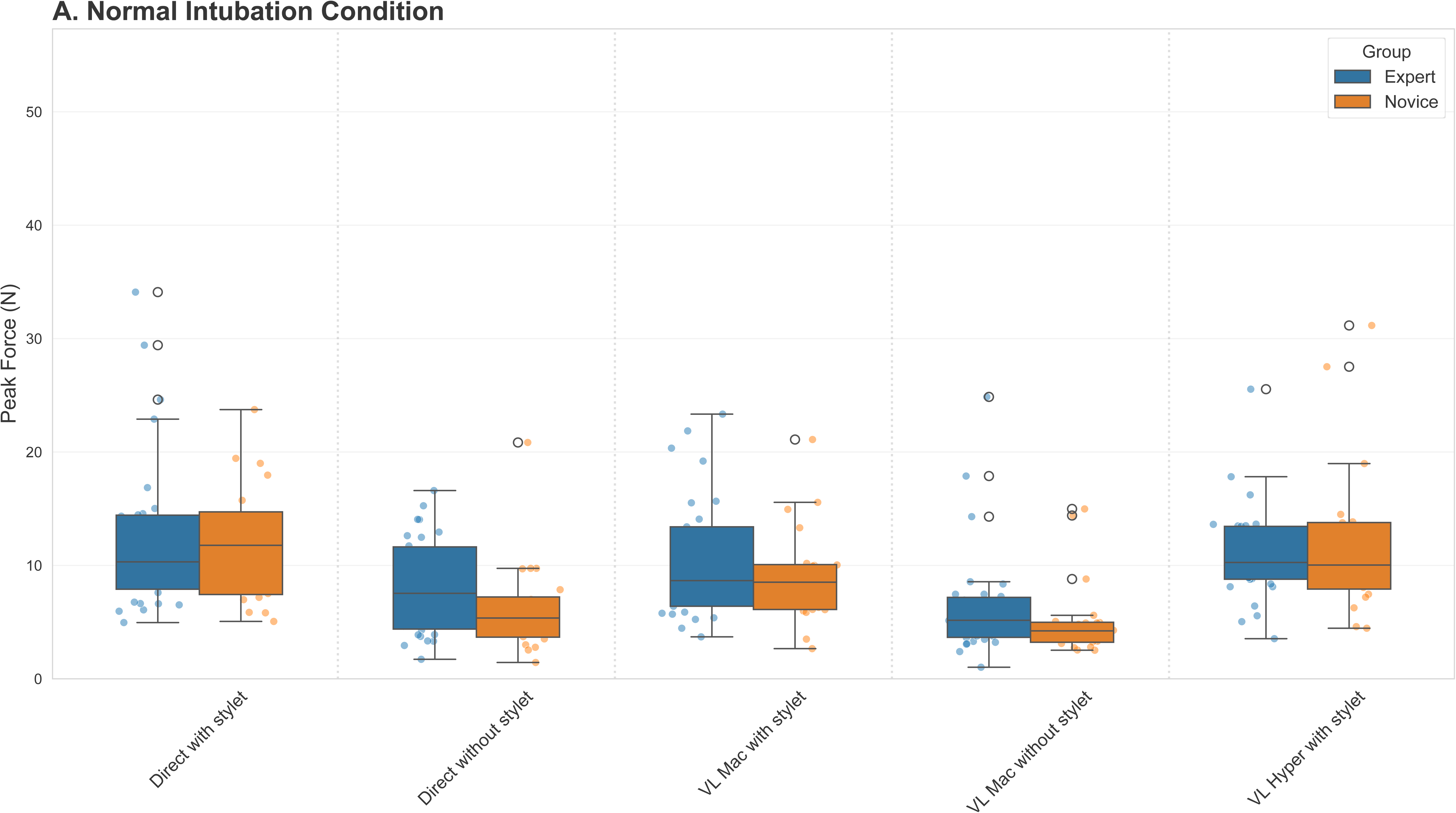
Summary figure showing peak glottic force distributions during normal condition intubation across techniques. Median and interquartile range are displayed for each technique–condition combination.

**Figure 3.**
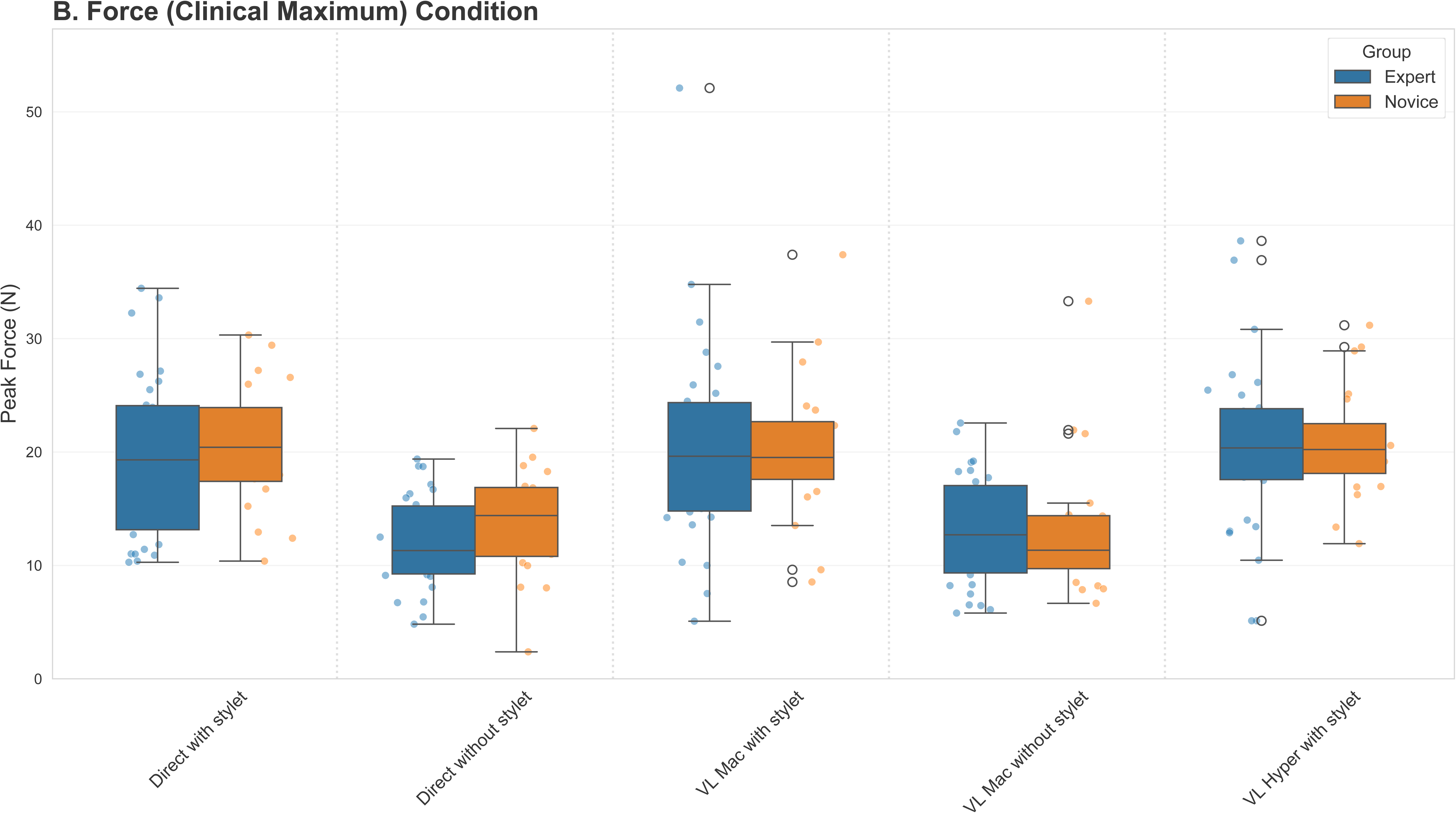
Summary figure showing peak glottic force distributions during simulated laryngospasm condition across techniques. Median and interquartile range are displayed for each technique–condition combination.

**Figure 4.**
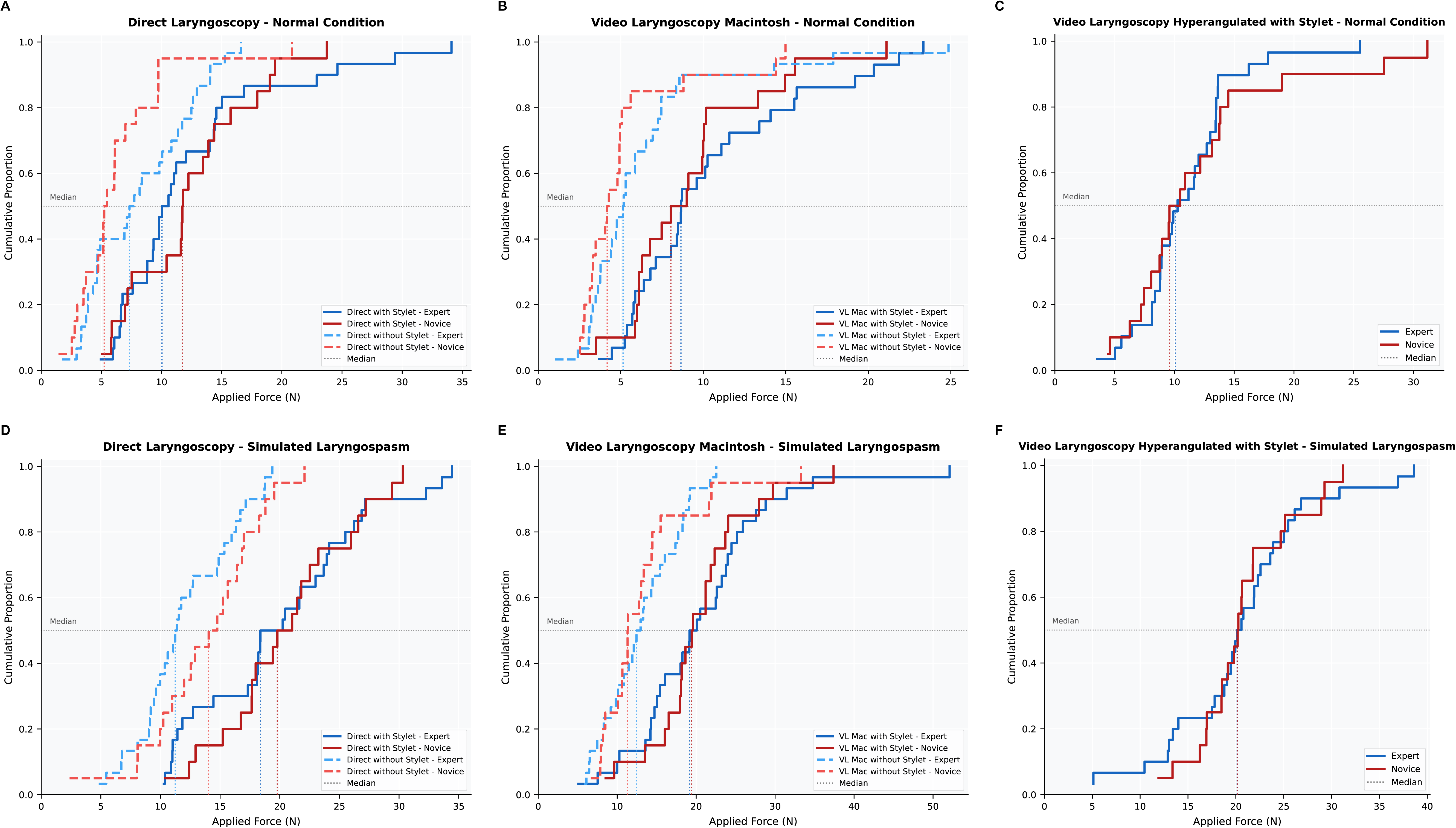
Cumulative distribution functions (CDFs) of the force applied to the glottis during endotracheal intubation across five techniques and two simulated clinical conditions, stratified by operator experience. Panels A–C depict the normal intubation condition and panels D–F depict the simulated laryngospasm condition, for direct laryngoscopy (A, D), video laryngoscopy with a Macintosh blade (B, E), and video laryngoscopy with a hyperangulated blade (C, F). Blue curves represent expert operators and red curves represent novice operators; solid lines indicate intubation with a stylet and dashed lines without a stylet (except for the hyperangulated blade, for which only the with-stylet configuration was tested). Vertical dotted lines indicate the median force for each group. Forces are expressed in Newtons (N).”

### Experienced Versus Less Experienced Operator Comparison

Mann-Whitney U tests comparing peak glottic forces between experienced and less experienced operators revealed no statistically significant differences for any technique–condition combination after Holm-Bonferroni correction (all adjusted p = 1.0; Table 3, Figures 2,3 and 4). Effect sizes were uniformly small to negligible, with rank-biserial correlations ranging from r = −0.27 to r = 0.27. The largest effect was observed for direct laryngoscopy without stylet under the standard condition, where experienced operators showed a numerically higher median force (7.5 N vs. 5.4 N; r = 0.27, p = 0.113), though this did not reach statistical significance after correction for multiple comparisons.

**Table 3.** Mann-Whitney U test results comparing peak glottic force between experienced and less experienced operators.

| Technique | Condition | n<br>Exp | n<br>Less | Median<br>Exp | Median<br>Less | U | r | p<br>(Holm) |
| --- | --- | --- | --- | --- | --- | --- | --- | --- |
| DL + stylet | Standard | 30 | 20 | 10.3 | 11.8 | 273.0 | 0.09 | 1.0 |
| DL + stylet | Clin. max | 30 | 20 | 19.3 | 20.4 | 283.5 | 0.06 | 1.0 |
| DL – stylet | Standard | 30 | 20 | 7.5 | 5.4 | 220.0 | 0.27 | 1.0 |
| DL – stylet | Clin. max | 30 | 20 | 11.3 | 14.4 | 221.0 | –0.26 | 1.0 |
| VL Mac + stylet | Standard | 29 | 20 | 8.7 | 8.5 | 258.0 | 0.11 | 1.0 |
| VL Mac + stylet | Clin. max | 30 | 20 | 19.6 | 19.5 | 299.0 | 0.0 | 1.0 |
| VL Mac – stylet | Standard | 30 | 20 | 5.2 | 4.2 | 230.5 | 0.23 | 1.0 |
| VL Mac – stylet | Clin. max | 30 | 20 | 12.7 | 11.3 | 282.5 | 0.06 | 1.0 |
| VL Hyper + stylet | Standard | 29 | 20 | 10.3 | 10.0 | 289.0 | 0.0 | 1.0 |
| VL Hyper + stylet | Clin. max | 30 | 20 | 20.4 | 20.2 | 295.5 | 0.02 | 1.0 |

*Exp, experienced; DL, direct laryngoscopy; VL Mac, videolaryngoscopy Macintosh; VL Hyper, videolaryngoscopy hyperangulated; r, rank-biserial correlation; clin. max, clinical maximum force. All p-values adjusted using Holm-Bonferroni method.*

### Effect of Stylet on Applied Force

The use of a stylet significantly increased the peak glottic force across all technique–condition combinations and in all groups (Table 4 and Figure 4 panels A, B, D and E). For the combined cohort under standard intubation conditions, the stylet increased the median force with direct laryngoscopy from 6.1 N (IQR: 3.9–9.8) to 11.1 N (IQR: 7.5–14.5), a median paired difference of 4.4 N (IQR: 2.0–8.7; p < 0.001, r = 0.80). For videolaryngoscopy, the stylet increased the median force from 4.8 N (IQR: 3.3–6.5) to 8.7 N (IQR: 6.1–12.4), with a median paired difference of 3.6 N (IQR: 1.2–7.2; p < 0.001, r = 0.88).

**Table 4.** Wilcoxon signed-rank test results for the effect of stylet use on peak glottic force (N).

| Group | Comparison | n | Median +stylet | Median –stylet | Median diff | r | p |
| --- | --- | --- | --- | --- | --- | --- | --- |
| Experienced | DL, standard | 30 | 10.3 | 7.5 | 3.4 | 0.75 | <0.001 |
| Experienced | DL, clin. max | 30 | 19.3 | 11.3 | 6.5 | 1.00 | <0.001 |
| Experienced | VL Mac, standard | 29 | 8.7 | 5.2 | 3.5 | 0.82 | <0.001 |
| Experienced | VL Mac, clin. max | 30 | 19.6 | 12.7 | 6.9 | 0.93 | <0.001 |
| Less exp. | DL, standard | 20 | 11.8 | 5.4 | 6.2 | 0.84 | 0.001 |
| Less exp. | DL, clin. max | 20 | 20.4 | 14.4 | 5.4 | 0.93 | <0.001 |
| Less exp. | VL Mac, standard | 20 | 8.5 | 4.2 | 3.9 | 0.93 | <0.001 |
| Less exp. | VL Mac, clin. max | 20 | 19.5 | 11.3 | 7.3 | 0.94 | <0.001 |
| All | DL, standard | 50 | 11.1 | 6.1 | 4.4 | 0.80 | <0.001 |
| All | DL, clin. max | 50 | 20.0 | 12.2 | 6.1 | 0.98 | <0.001 |
| All | VL Mac, standard | 49 | 8.7 | 4.8 | 3.6 | 0.88 | <0.001 |
| All | VL Mac, clin. max | 50 | 19.5 | 12.3 | 7.0 | 0.94 | <0.001 |

*VL Mac, videolaryngoscopy Macintosh; diff, median of paired differences; r, matched-pairs rank-biserial correlation; clin. max, clinical maximum force. All p-values are Holm-Bonferroni adjusted.*

Under the simulated laryngospasm condition, the stylet effect was even more pronounced. Direct laryngoscopy force increased from 12.2 N to 20.0 N (median difference 6.1 N, IQR: 2.4–10.8; p < 0.001, r = 0.98), and videolaryngoscopy force increased from 12.3 N to 19.5 N (median difference 7.0 N, IQR: 3.0–10.5; p < 0.001, r = 0.94). All comparisons yielded large effect sizes (rank-biserial r ≥ 0.75), indicating a robust and clinically meaningful increase in glottic force attributable to stylet use.

### Comparison Across Laryngoscopy Techniques

The Friedman test for repeated measures comparing forces across the three techniques using a stylet (direct laryngoscopy, videolaryngoscopy Macintosh, and videolaryngoscopy hyperangulated) revealed a significant difference among techniques under the standard intubation condition for the combined cohort (χ² = 6.98, df = 2, p = 0.030; Table 5). Post-hoc Conover pairwise comparisons with Holm-Bonferroni correction revealed that videolaryngoscopy Macintosh produced significantly lower forces than hyperangulated videolaryngoscopy (median 8.7 N vs. 10.3 N; mean rank 1.69 vs. 2.18; adjusted p = 0.049). The difference between videolaryngoscopy Macintosh and direct laryngoscopy showed a trend toward significance (adjusted p = 0.072). In the less experienced subgroup, a significant Friedman test was also observed (χ² = 6.10, p = 0.046), though no individual pairwise comparison reached significance after correction. In the experienced subgroup, no significant difference among techniques was observed under standard conditions (χ² = 1.93, p = 0.383). Under the simulated laryngospasm condition, no significant differences among techniques were found in any group (all p > 0.50).

**Table 5.** Friedman test results comparing peak glottic force across three techniques (with stylet).

| Group | Condition | n | DL Median<br>(rank) | VL Mac Median<br>(rank) | VL Hyper Median<br>(rank) | $\chi^2$ | p |
| --- | --- | --- | --- | --- | --- | --- | --- |
| Experienced | Standard | 29 | 10.0 (2.07) | 8.7 (1.79) | 10.3 (2.14) | 1.93 | 0.383 |
| Less exp. | Standard | 20 | 11.8 (2.20) | 8.5 (1.55) | 10.0 (2.25) | 6.10 | 0.046* |
| All | Standard | 49 | 11.0 (2.12) | 8.7 (1.69) | 10.3 (2.18) | 6.98 | 0.030* |
| Experienced | Clin. max | 30 | 19.3 (2.00) | 19.6 (1.93) | 20.4 (2.07) | 0.27 | 0.872 |
| Less exp. | Clin. max | 20 | 20.4 (1.95) | 19.5 (1.85) | 20.2 (2.20) | 1.30 | 0.527 |
| All | Clin. max | 50 | 20.0 (1.98) | 19.5 (1.90) | 20.2 (2.12) | 1.24 | 0.543 |

\* $p < 0.05$ . DL, direct laryngoscopy; VL Mac, videolaryngoscopy Macintosh; VL Hyper, videolaryngoscopy hyperangulated; $\chi^2$ , Friedman chi-squared statistic; clin. max, clinical maximum force. Median values in Newtons; mean ranks in parentheses

#### Anthropometric Correlations

Spearman rank correlation analysis revealed no significant correlations between grip strength or hand size and the peak glottic force applied during any technique–condition combination (Table 6). Correlation coefficients for grip strength ranged from ρ = −0.26 (direct laryngoscopy with stylet, standard condition) to ρ = 0.17, with all p-values > 0.05. The overall average force across all conditions was not correlated with either grip strength (ρ = −0.09, p = 0.541) or hand size (ρ = −0.16, p = 0.252).

**Table 6.**
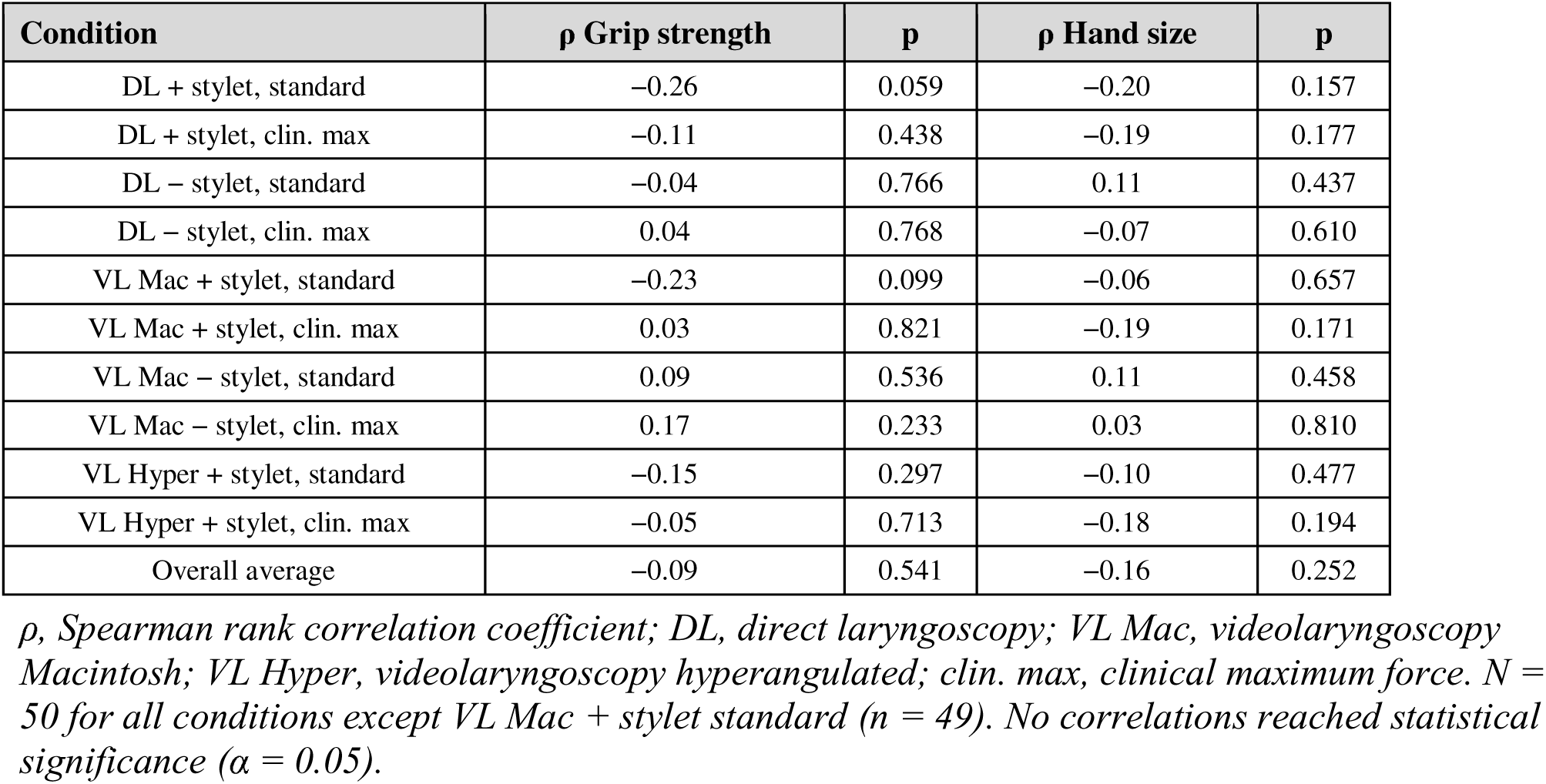
Spearman rank correlation coefficients between anthropometric variables and peak glottic force.

| Condition | $\rho$ Grip strength | p | $\rho$ Hand size | p |
| --- | --- | --- | --- | --- |
| DL + stylet, standard | -0.26 | 0.059 | -0.20 | 0.157 |
| DL + stylet, clin. max | -0.11 | 0.438 | -0.19 | 0.177 |
| DL – stylet, standard | -0.04 | 0.766 | 0.11 | 0.437 |
| DL – stylet, clin. max | 0.04 | 0.768 | -0.07 | 0.610 |
| VL Mac + stylet, standard | -0.23 | 0.099 | -0.06 | 0.657 |
| VL Mac + stylet, clin. max | 0.03 | 0.821 | -0.19 | 0.171 |
| VL Mac – stylet, standard | 0.09 | 0.536 | 0.11 | 0.458 |
| VL Mac – stylet, clin. max | 0.17 | 0.233 | 0.03 | 0.810 |
| VL Hyper + stylet, standard | -0.15 | 0.297 | -0.10 | 0.477 |
| VL Hyper + stylet, clin. max | -0.05 | 0.713 | -0.18 | 0.194 |
| Overall average | -0.09 | 0.541 | -0.16 | 0.252 |

$\rho$ , Spearman rank correlation coefficient; DL, direct laryngoscopy; VL Mac, videolaryngoscopy Macintosh; VL Hyper, videolaryngoscopy hyperangulated; clin. max, clinical maximum force. $N = 50$ for all conditions except VL Mac + stylet standard ( $n = 49$ ). No correlations reached statistical significance ( $\alpha = 0.05$ ).

## DISCUSSION

The principal findings of this study are as follows: (1) Under standard intubation conditions, the median force applied to the glottis ranged from 4.8 to 11.1 N and from 12.3 to 20.2 under simulated laryngospasm conditions. (2) No significant difference in glottic force was observed between experienced and less experienced operators for any intubation technique or condition. (3) The use of a stylet consistently and significantly increased the force applied on the glottis, with median increases of 3.4 to 7.3 N and uniformly large effect sizes (r ≥ 0.75), regardless of technique, condition, or experience level. (4) Under standard intubation conditions, videolaryngoscopy with a Macintosh blade produced the lowest glottic force among the three techniques, with a significant difference compared to hyperangulated videolaryngoscopy (p = 0.049). This advantage was not maintained under the simulated laryngospasm condition. (5) Neither hand grip strength nor hand size was correlated with the force applied during intubation.

### Force Application on the glottis

Our results provide the first comprehensive quantitative characterization of the force transmitted to the glottis during endotracheal intubation — specifically the force conveyed by the right hand through the tube and stylet. We examined five intubation techniques across two simulated clinical scenarios. Under standard intubation conditions, the median glottic force ranged from 4.8 to 11.1 N, rising to 12.3 to 20.2 N under simulated laryngospasm. Even among experienced operators under maximal force conditions, the median glottic force did not exceed 31.1 N. Despite meaningful differences across techniques, the force applied to the glottis via the tube appears overall lower than both the lifting force exerted by the laryngoscope — reported between 10 and 48.8 N depending on the device^8,9^ — and the forces associated with the cricoid pressure (30 to 44 N)^23–25^.

### Experience Level and Force Application

The absence of any significant difference in applied glottic force between experienced and less experienced operators, across all ten technique–condition combinations, contrasts with our initial hypothesis and the commonly held assumption that less experienced operators apply excessive force during intubation. Despite substantial differences in clinical experience (76.7% of experienced operators performing >50 intubations per 3-month period vs. 60% of less experienced operators performing 0–5), both groups applied remarkably similar forces, with uniformly negligible to small effect sizes (|r| ≤ 0.27).

While the literature clearly demonstrates that less experienced operators have lower first-pass success rates and higher complication rates^26^, our results suggest that these adverse outcomes may not be attributable to differences in applied force per se. Instead, difficulties may stem from suboptimal laryngoscope positioning, inadequate glottic visualization, or less efficient tube delivery. Zamora et al. similarly reported inconsistent force differences between experienced and less experienced operators despite clear differences in technique metrics^27^. Sakles et al. have shown that learning curves for intubation are primarily defined by visualization and success metrics rather than force control.^12^

From a clinical perspective, if force application is not a distinguishing feature between experience levels, teaching strategies focused on reducing force may be less impactful than approaches targeting visualization optimization and tube delivery technique. However, our study was conducted on a manikin model where anatomical variability and tissue compliance are standardized; in live patients with variable airway anatomy and pathology, experience-related differences in force application may emerge.

### The Stylet Effect: A Robust Increase in Glottic Force

The most striking finding of this study is the consistent and significant increase in glottic force associated with stylet use. Across all groups and conditions, the stylet increased the median peak force by 3.4 to 7.3 N, with uniformly large effect sizes (rank-biserial r ≥ 0.75). Under standard conditions, the stylet nearly doubled the median force for direct laryngoscopy (from 6.1 to 11.1 N) and videolaryngoscopy (from 4.8 to 8.7 N). This effect was observed in both experience groups, indicating that the force increase is an inherent biomechanical consequence of tube rigidity rather than a reflection of operator skill.

These results should be interpreted in the context of the established benefits of stylet use. The STYLETO trial demonstrated that the stylet significantly improves first-attempt intubation success (78.2% vs. 71.5%, p = 0.01) ^11^. The increased force observed in our study must therefore be weighed against the reduced risk of multiple intubation attempts, which themselves carry substantial risk of airway trauma. Strategies to mitigate the force increase, such as partial stylet withdrawal after the tube tip passes the vocal cords, use of malleable rather than rigid stylets, and careful attention to stylet tip positioning, are recommended to optimize the risk–benefit balance^10,28^.

### Technique Comparison: The Advantage of Videolaryngoscopy

Under standard intubation conditions, videolaryngoscopy with a Macintosh blade produced the lowest forces among the three techniques compared (all using a stylet), with a significant Friedman test for the combined cohort (χ² = 6.98, p = 0.030) and a significant post-hoc difference compared to hyperangulated videolaryngoscopy (adjusted p = 0.049). Previous studies have demonstrated that videolaryngoscopy reduces the lifting force applied by the left hand.^8,9^ Our findings extend this observation by showing that videolaryngoscopy with a Macintosh blade also results in lower glottic contact forces from the right-hand during tube passage.

The finding that hyperangulated videolaryngoscopy produced higher glottic forces than Macintosh videolaryngoscopy during standard intubation may seem counterintuitive, given that hyperangulated blades are generally associated with reduced lifting force for glottic exposure. However, hyperangulated blades require a correspondingly shaped stylet and a more complex tube delivery trajectory, which may result in greater contact forces at the glottis during tube advancement^29^. This distinction is clinically important: while hyperangulated blades may reduce the force of laryngoscopy itself, they may increase the force of tube delivery through the glottis.

Importantly, the force advantage of videolaryngoscopy was abolished under the simulated laryngospasm condition, where all three techniques yielded similar forces (approximately 19–20 N median). This suggests that the technique-related differences observed during standard intubation reflect the inherent biomechanical requirements of each device for tube delivery, rather than a fixed ceiling effect.

### Anthropometric Characteristics and Intubation Force

Neither grip strength nor hand size was significantly correlated with applied force in any technique–condition combination. The forces involved in endotracheal intubation (5–31 N in our study) are well within the capability range of all operators regardless of grip strength (median 33–41 kg = 323–402 N), indicating that intubation force is not limited by physical capacity but rather determined by technique and equipment interactions with the airway. Concerns that practitioners with smaller hands or lower grip strength may apply different forces during intubation are not supported by our data.

### Limitations

Several limitations should be acknowledged. First, this study was conducted using a manikin model. While this design allowed standardized and reproducible conditions, manikin airways do not fully replicate the compliance, tissue properties, and anatomical variability of human airways. The forces measured may differ from those encountered in clinical practice, particularly in patients with difficult airway anatomy, obesity, or cervical pathology.

Second, the force measurement was limited to peak force at the glottic level. Other force metrics, such as impulse force (the integral of force over time) and the distribution of force across different airway structures, may provide additional insights into the mechanisms of airway trauma. Cordovani et al. demonstrated that while peak forces were lower with the GlideScope, impulse forces were similar due to longer laryngoscopy duration.^9^

Third, the sample size was based on convenience sampling reflecting operator availability at a single centre. The uniformly small effect sizes observed (|r| ≤ 0.27) suggest that any true difference, if present, would be of limited clinical significance. Fourth, the less experienced group encompassed a heterogeneous population including respiratory therapists, medical students, and general surgeons with varying levels of exposure to intubation. Fifth, the physical maximum force condition was performed only by a subset of experienced operators (n = 10), limiting the generalizability of this condition. Finally, while we randomized technique order, we did not control for other factors that may influence applied force, such as fatigue or familiarity with specific laryngoscope models.

## CONCLUSION

This study demonstrates that the force applied on the glottis during endotracheal intubation is primarily determined by the intubation technique and the use of a stylet, rather than by operator experience or anthropometric characteristics. The use of a stylet consistently and substantially increased glottic force (representing the single largest modifiable factor influencing applied force. Under standard intubation conditions, videolaryngoscopy with a Macintosh blade produced the lowest forces among the three techniques, with a significant advantage over hyperangulated videolaryngoscopy. No significant differences were observed between experienced and less experienced operators, and no correlations were found between operator grip strength, hand size, and intubation force.

Future studies should validate these findings in clinical settings with live patients, incorporate additional force metrics (impulse force, pressure distribution), and explore whether the force differences observed translate into measurable differences in airway morbidity. The development of force-sensing laryngoscopes for routine clinical use could enable real-time force feedback during intubation, potentially reducing airway trauma and improving training outcomes.

## AUTHORSHIP ATTESTATIONS

**Louis Morisson:** conception and design, statistical analysis, drafting of the manuscript and final approval of the version to be published.

**Arnaud Latreille:** data acquisition and final approval of the version to be published

**Mathias Pietrancosta:** data acquisition, critical revision of the manuscript and final approval of the version to be published.

**Katia Djerroud:** statistical analysis and final approval of the version to be published

**Issam Tanoubi:** conception and design, critical revision of the manuscript and final approval of the version to be published.

**Thomas Hemmerling:** conception and design, critical revision of the manuscript and final approval of the version to be published.

**Pascal Laferrière-Langlois:** conception and design, data acquisition, statistical analysis, drafting and critical revision of the manuscript and final approval of the version to be published.

## Data Availability

All data produced in the present study are available upon reasonable request to the authors

## ACKNOWLEDGEMENTS

Dr. Louis Morisson is supported by the Canadian Institutes of Health Research (CIHR, Canada, 199277) and the Fonds de recherche du Québec (FRQ, Canada, https://doi.org/10.69777/369069). Pascal Laferrière-Langlois is supported by the Fonds de recherche du Québec (FRQ, Canada).

